# Uterine fibroids and risk of hypertensive disorders of pregnancy – results from a racially diverse high-risk cohort

**DOI:** 10.1101/2024.03.05.24303830

**Authors:** K. Cameron, M. Borahay, X Hong, V. Baker, A. Vaught, X. Wang

## Abstract

**Study Question:** What is the impact of the presence of uterine fibroids on the risk of developing hypertensive disorders of pregnancy (HDP) in a predominantly urban, low-income, Black, and Hispanic population of women with ultrasound or clinically diagnosed uterine fibroids with rich phenotypic data to carefully control for potential confounders?

**Summary answers:** The odds of HDP were 39% higher in women with uterine fibroids compared to those without when controlled for age at delivery, race, prepregnancy BMI, education, parity, and smoking status; neither fibroid location or size modified this risk.

**What is known already:** Studies are conflicting regarding the impact of uterine fibroids on risk of HDP; limitations of prior studies include primarily Western European populations and lack of measurement of potential confounders.

**Study design, size, and duration:** A total of 7030 women from the Boston Birth Cohort (a racially diverse cohort recruited from 1998 to 2018) that had clinical and ultrasound data regarding uterine fibroid status were included in this analysis.

**Participants/materials, setting, and methods:** Four hundred eighty-nine women with uterine fibroids and 6541 women without were included. Hypertensive disorders of pregnancy were ascertained from medical records. Logistic regression was performed to assess the risk of HDP in women with and without uterine fibroids. Covariates adjusted for included age at delivery, race, pre-pregnancy BMI, education, parity, and smoking status during pregnancy. Sub-analyses were performed to assess the impact of specific fibroid location and overall fibroid volume burden.

**Main results and the role of chance:** The incidence of uterine fibroids in the cohort was 7% (N=489). Twelve percent of women without uterine fibroids and 17% of women with fibroids developed HDP; in multivariate analyses adjusted for the potential confounders above, the odds of HDP were 39% higher in women with uterine fibroids compared to those without (p=0.03). Women with a uterine fibroid diagnosis based on ICD code (n=297) versus asymptomatic incidental ultrasound diagnosis (n=192) had a significantly greater chance of developing HDP (20 vs 15%, p=0.006). There did not appear to be an association between number of fibroids or total fibroid volume and the risk of developing HDP. *Limitations, reasons for caution:* This study has a relatively small sample size. While post-hoc power calculation determined that there was adequate power to detect a 4.6% difference in the incidence of development of HDP between participants with uterine fibroids and those without, the sub-analyses based on fibroid size, location, and method of diagnosis were underpowered to determine a similar level of difference.

**Wider implications of the findings:** In a racially diverse cohort, presence of uterine fibroids was a significant risk factor for developing HDP, regardless of uterine fibroid size or location. This may have implications for additional monitoring and risk stratification in women with uterine fibroids.

**Study funding/competing interests:** KC supported by WRHR NIH NICHD Award # K12 HD103036, PI Andrew Satin, RD James Segars. The Boston Birth Cohort (the parent study) was supported in part by the National Institutes of Health (NIH) grants (2R01HD041702, R01HD098232, R01ES031272, R01ES031521, and U01 ES034983); and the Health Resources and Services Administration (HRSA) of the U.S. Department of Health and Human Services (HHS) (UT7MC45949). This information or content and conclusions are those of the authors and should not be construed as the official position or policy of, nor should any endorsements be inferred by any funding agencies.

**Trial registration number:** The BBC is registered under clinicaltrials.gov NCT03228875.

## Introduction

Uterine leiomyomas (fibroids) are the most common solid symptomatic neoplasm in women, estimated to occur in up to 70% of women by the time of menopause (Stewart and Borah, 2021). Increasing data support an association between uterine fibroids and development of chronic hypertension over the life course (Brewster *et al*., 2022, Haan *et al*., 2018). Since pregnancy may be viewed as a “cardiac stress test”, with the development of preeclampsia in pregnancy linked to the development of cardiovascular disease long-term (Ying *et al*., 2018), understanding the association between uterine fibroids and hypertensive disorders in pregnancy (HDP) has important implications for both offspring and maternal health. While obstetrical complications resulting from the presence of uterine fibroids, including spontaneous abortion, fetal malpresentation, preterm labor, postpartum hemorrhage, and cesarean section have been well-described (Stout *et al*., 2010) (Michels *et al*., 2014) (Coronado *et al*., 2000), the relationship between uterine fibroids and development of HDP has been conflicting. Prior studies have been limited by primarily western European or Asian cohorts and a lack of measurement of important potential confounders including shared risk factors like body mass index (BMI) and race (Biderman-Madar *et al*., 2005) (Girault *et al*., 2018) (Conti *et al*., 2013) (Harlev *et al*., 2019). Fibroids are more prevalent and occur at younger ages in black women, who also have a higher rate of maternal morbidity and mortality; given these limitations, The Proceedings from the Third National Institutes of Health International Congress on Advances in Uterine Leiomyoma Research has called on researchers to diversify study populations of fibroid patients (Segars *et al*., 2014). Therefore, the goals of this study were to use a birth cohort of racially diverse participants with well-phenotyped uterine fibroids to assess the impact of the presence of uterine fibroids on the risk of developing hypertensive disorders of pregnancy.

## Materials and Methods

### Study population

The study population is a subset of the Boston Birth Cohort (BBC), an ongoing longitudinal cohort study begun in 1998 at the Boston Medical Center in Massachusetts. Detailed information about the BBC has been reported elsewhere (Pearson *et al*., 2022). Briefly, mothers who delivered singleton live births were eligible for the study and were invited to participate within 1 to 3 days after delivery. Pregnancies that were multiple gestations, those with fetal chromosomal abnormalities or major birth defects, preterm deliveries (PTD) due to non-obstetric factors (such as trauma), and pregnancies that were a result of in vitro fertilization were not included. The BBC is a low-income patient population, with a relatively high proportion of PTD and HDP. After obtaining written informed consent, each mother was interviewed using a standardized questionnaire to gather dietary and epidemiologic data, and their electronic medical records (EMRs) were abstracted in completion. The study protocol was approved by the Institutional Review Boards of Boston Medical Center and the Johns Hopkins Bloomberg School of Public Health.

### Exposure

Uterine fibroids were identified through both use of International Classification of Diseases (ICD) 9 or 10 prior to and during pregnancy (See Supplementary Table 1 for diagnoses included) and through review of any radiologic ultrasound performed prior to and in pregnancy. Ultrasound reports with information regarding uterine fibroid size and location were abstracted for analysis. Several assumptions were made to allow for assessment of ultrasound reports. A uterine fibroid reported qualitatively as ‘small’ but without dimensions provided was recorded as 2cm, and a uterine fibroid reported qualitatively as ‘large’ but not measured was recorded as 4cm, based on the median fibroid size from the ultrasounds that had both quantitative measurements and qualitative descriptors. For calculating the volume of a uterine fibroid, if only a single measurement was provided this was utilized for the volume. For individuals with multiple ultrasound data points available, measurements from the ultrasound that was the closest to the date of delivery (pre-delivery) was utilized, with the exception of when multiple ultrasounds were performed within six months of one another and one included quantitative measurements and the other did not – in that case data from the ultrasound with quantitative data was utilized, even if it was not the closest to delivery.

### Outcome

Diagnoses of HDP (preeclampsia, eclampsia, and HELLP [hemolysis, elevated liver enzymes, and low platelet count] syndrome, superimposed preeclampsia and gestational hypertension), our outcome of interest, were based on physician diagnosis and manually abstracted from the maternal prenatal charts by trained research staff. Preeclampsia was defined, at the time, based on the National High Blood Pressure Education Program Working Group on High Blood Pressure in Pregnancy as systolic blood pressure of at least 140 mm Hg or diastolic blood pressure of at least 90 mm Hg; proteinuria of at least 1+ on at least 2 occasions with onset after 20 weeks of gestation; or worsening chronic hypertension (systolic blood pressure, ≥160 mm Hg; diastolic blood pressure, ≥110 mm Hg). Physician-diagnosed preeclampsia, eclampsia, HELLP, gestational hypertension, and superimposed preeclampsia were further confirmed by review of all relevant medical records to meet the definitions according to the recent American College of Obstetricians and Gynecologist (ACOG) criteria (Anonymous, 2019).

### Other covariates

Gestational age was assessed by early prenatal ultrasound (<L20Lweeks) or based on the first day of the last menstrual period as recorded in maternal EMRs if early prenatal ultrasound was not available. Using a standard maternal questionnaire interview(Pearson *et al*., 2022), maternal epidemiological factors were collected including race/ethnicity, maternal age at delivery, highest education level, parity, smoking during pregnancy, alcohol drinking during pregnancy, illicit drug use during pregnancy, lifetime stress, previous history of PTD, and maternal birthplace (US-born/non-US-born). Maternal pregestational body mass index (BMI) was calculated as self-reported weight (kg) divided by height squared (m2). Clinical complications before pregnancy, including chronic hypertension and pregestational diabetes, were defined based on archived EMRs.

## Results

A total of 7,030 participants were included in this analysis. Demographic characteristics for included participants with and without uterine fibroids are summarized in Table 1. In addition, demographic characteristics for participants based on method of uterine fibroid diagnosis (ICD, radiologic review, or both) are included in Supplementary Table 2. A total of 7% of women in the Boston Birth Cohort had a diagnosis of uterine fibroids. Twelve percent of women without uterine fibroids developed HDP, and seventeen percent of women with uterine fibroids developed HDP (p=0.001 in univariate analysis). In multivariate analyses controlled for age at delivery, race, pre-pregnancy BMI, education, parity, and smoking status, the odds of HDP were 39% higher in women with uterine fibroids compared to those without (p=0.03). Furthermore, when restricted only to analysis of Black women (N=4454), the odds of HDP were 50% higher in women with uterine fibroids compared to those without when controlled for the same covariates, and when restricted to Hispanic women (N=2440) the odds of HDP were 54% higher in women with uterine fibroids compared to those without when controlled for age at delivery, pre-pregnancy BMI, education, parity, and smoking status.

**Table 1.** Demographic Characteristics of nested cohort mother-child dyads, BBC.

|  |  |  |
| --- | --- | --- |
| **Maternal demographic characteristics at enrollment of pregnancy** | **Unaffected controls**<br>**N=6,541** | **Uterine fibroids**<br>**N=489** |
| Maternal age at delivery(year), Mean (SD) | 27.9 (6.5) | 33.1 (5.6) ** |
| Race/Ethnicity |  |  |
| Black, % | 50 | 75 ** |
| White, % | 12 | 5 |
| Hispanic, % | 29 | 14 |
| BMI, Mean (SD) | 25.8 (6.1) | 28.3 (7.2) ** |
| Nulliparity, % | 41 | 38 |
| Maternal education< college, % | 67 | 54 ** |
| Maternal ever smoking during pregnancy, % | 19 | 13 ** |
| Gestational age at delivery(weeks), Mean (SD) | 38.0 (3.1) | 37.0 (3.7) ** |
| Preterm delivery <37 weeks, % | 26 | 37 ** |
| Birthweight (grams), Mean (SD) | 2,967 (760) | 2,822 (870) |
| Mode of delivery |  |  |
| Cesarean section, % | 31 | 53 ** |
| Vaginal delivery, % | 69 | 47 |
| Hypertensive Disorders of Pregnancy, % | 12 | 19 ** |
| Gestational diabetes, % | 5 | 7 |
| ** p<0.05 compared to unaffected controls in unadjusted univariate analysis |  |  |

Sub-analyses were performed based on method of uterine fibroid diagnosis, given that uterine fibroids are often underdiagnosed and symptomatic uterine fibroids versus incidentally noted uterine fibroids may have different risk profiles. Women with a uterine fibroid diagnosed based on ICD code (n=297) versus asymptomatic incidental ultrasound diagnosis without accompanying ICD diagnosis (n=192) had a significantly greater chance of developing HDP (20 vs 15%, p=0.006); however, even in those with incidentally noted uterine fibroid(s), the odds of HDP were 18% higher in women compared to those without uterine fibroids (p=0.048).

The degree of uterine fibroid burden impacting risk was also analyzed. For the women with ultrasound imaging noting uterine fibroids, the median total number of fibroids was 2 (range 1-6). The mean volume of the total fibroid burden was 3.67cm^3^ (95% CI 1.78 – 13.7cm^3^). The mean volume of the largest fibroid was 3.1cm^3^ (95% CI 1.6 - 9.1cm^3^). Ten percent of women had at least one submucosal fibroid, 47% had an intramural fibroid, and 43% had a subserosal uterine fibroid. In logistic regression models, neither the impact of the size of the largest fibroid, the total fibroid volume, nor the presence of a submucosal fibroid compared to other location mediated the risk of developing HDP.

Finally, women with pre-existing chronic hypertension were censored from the dataset (N=464 women total, 64 with fibroids and 300 without) and this did not chance the magnitude of the effect of odds of HDP based on fibroid presence (p=0.61).

## Discussion

Uterine fibroids are highly prevalent tumors in reproductive-aged women, and they pose a significant health burden that disproportionally effects Black women(Kjerulff *et al*., 1996). In addition to their debilitating symptoms and impact on quality of life(Stewart and Borah, 2021), uterine fibroids are now increasingly associated with the long-term risk of developing cardiovascular disease(Brewster *et al*., 2022, Haan *et al*., 2018, Uimari *et al*., 2016), though whether that is due to shared risk factors or underlying pathophysiology is largely unknown. Given that uterine fibroids will impact up to 10% of all pregnancies(Zhao *et al*., 2020), there is a critical need to understand the impact of uterine fibroids on hypertensive disorders in pregnancy, as this can have a significant impact on maternal and offspring long-term health(Alsnes *et al*., 2017, Boyd *et al*., 2017, Nahum Sacks *et al*., 2018).

Prior studies regarding the presence of uterine fibroids and the risk of HDP have been conflicting with some studies reporting an association and others not finding any impact of the presence of uterine fibroid. Some of the largest studies to examine this question have been conducted in Asian countries without inclusion of women of African descent (Chen *et al*., 2021) (Lee *et al*., 2020) (Gong *et al*., 2022) (Pan *et al*., 2019), while others have focused only on infertile women requiring fertility treatment (Biderman-Madar *et al*., 2005), or have utilized only ICD diagnosis without imaging (Farland *et al*., 2019), or imaging only in the second trimester of pregnancy when fibroids may be underreported (Farland *et al*., 2019, Stout *et al*., 2010). Furthermore, collection of important shared risk factors including BMI and participant age were not uniformly reported.

Our study found that the presence of uterine fibroids was associated with obstetrical outcomes already well-established to be impacted by fibroids, including pre-term birth and delivery via cesarean section (Table 1), confirming the reliability of our cohort. In addition, we found that traditional risk factor associations with HDP were present in our cohort (increased risk with increasing age and BMI, inverse relationship with parity and smoking status, Table 2). We found that even when controlled for important shared risk factors, including age, BMI, and parity the presence of uterine fibroids was independently associated with a significantly increased risk of developing HDP. This risk was also independent of the size, total volume burden, or location of the uterine fibroids. This may indicate that the systemic angiogenic and vasogenic changes that promote fibroid development, rather than the mass effect of the fibroid itself, play a role in abnormal placentation and HDP disease process.

**Table 2.** Logistic Regression Analysis of Risk of HDP.

| **Table 2. Logistic Regression Analysis of Risk of HDP** |  |  |  |  |  |
| --- | --- | --- | --- | --- | --- |
| Variable | Odds Ratio | Standard Error | P-value | 95% CI |  |
| Uterine fibroids (y/n) | 1.39 | 0.21 | **0.03** | 1.03 | 1.88 |
| Maternal age (continuous) | 1.02 | 0.01 | **0.02** | 1.00 | 1.03 |
| Race (ref=Black, 1=Black) | 1.01 | 0.07 | 0.18 | 0.94 | 1.33 |
| BMI (continuous) | 1.05 | 0.01 | **<0.01** | 1.04 | 1.07 |
| Education (ref=less than college, 1=college or above) | 0.99 | 0.04 | 0.86 | 0.92 | 1.07 |
| Parity (ref=nulliparous, 1=P1 or greater) | 0.54 | 0.05 | **<0.01** | 0.45 | 0.66 |
| Smoking 6 months prior to conception or in pregnancy (ref=never, 1=ever) | 0.78 | 0.09 | **0.03** | 0.62 | 0.98 |
| Intercept | 0.03 | 0.01 | **<0.01** | 0.01 | 0.05 |

Importantly, the degree of risk that the presence of uterine fibroid(s) imparts was more pronounced in African American and Hispanic women in analyses controlled for shared risk factors like age and BMI. Furthermore, while women with an ICD diagnosis, indicating clinically symptomatic disease, had a greater risk than women with uterine fibroids noted incidentally on imaging, even women with incidentally noted uterine fibroids had a significantly greater risk of developing HDP than women without uterine fibroids.

The two largest studies to previously examine the relationship between uterine fibroids and HDP in the United States include Stout et al and Farland et al (Farland *et al*., 2019, Stout *et al*., 2010). Stout et al utilized second trimester fetal anatomy ultrasounds to examine the impact of uterine fibroids on deliveries from 1990-2007 in a single center in St. Louis, Missouri. The prevalence of uterine fibroids in this population was 3.2% and only approximately 20% of the study population was African American; no data are reported for Hispanic recipients. This study did not demonstrate an increase in preeclampsia for women with uterine fibroids in models adjusting for similar covariates to our study, though the p-value was 0.05. Farland et al examined over 92,000 pregnancies linked to hospital discharge records in the Massachussetts All Payers Claims Database. Uterine fibroids were identified by ICD diagnosis and the prevalence was 4.6%; 3.4% of the study population were Non-Hispanic Black women and 4.6% were Hispanic. In this study, the presence of uterine fibroids was associated with an increased relative risk of 12% for developing HDP. In neither study was sub-analysis for race performed.

Strengths of our study include the use of both ultrasound and ICD diagnoses, as this allowed for exploration of both overtly symptomatic disease and those with clinically ‘insignificant’ or overlooked fibroid disease. While an association with increased risk of uterine fibroids was seen for both groups, the risk was higher for clinically diagnosed uterine fibroids, indicating that potentially associated symptoms including presence of anemia may play a mediating role; this should be investigated in future studies. In addition, the presence of ultrasound imaging results in our study allowed for an assessment of the impact of fibroid burden and location.

The fact that the presence of uterine fibroids was associated with HDP, independent of size or location, is intriguing. There is some evidence that there may be a differential response to hypoxia in uterine fibroids compared to myometrial cells (Miyashita-Ishiwata *et al*., 2022), which could play a role in the pathogenesis of HDP. In addition, there is data from translational work that indicate that inflammation, alterations in the endometrial structure, and molecular signaling could all be postulated to contribute to a mechanism (Boynton-Jarrett *et al*., 2005, Humphrey, 2008, Kuwahara *et al*., 2002).

There are important limitations of this work. First, the use of this birth cohort by definition requires that a live birth occurred, and so this study was not able to assess the impact of the presence of uterine fibroids on previable delivery or intrauterine fetal death. Secondly, it is possible that some participants may have had uterine fibroids present on ultrasound imaging but it was not noted in the radiologic reports. Access to the images from the reports was not available at the time of this analysis. However, if there was an increased incidence of uterine fibroids in the group characterized without fibroids, that would only serve to bias our findings towards the null. In addition, this study population is derived from an urban, high risk catchment hospital with significant racial and ethnic diversity. While this may limit generalizability to low-risk populations, this is precisely the group of women for whom studies regarding the impact of uterine fibroids across the life course are needed.

Given the increased prevalence of HDP based on uterine fibroid status in this high-risk population, particular attention to individualized counseling and potentially appropriate clinical interventions and increased monitoring may be beneficial in individuals with this common uterine pathology, whether they are symptomatic or incidentally noted at the time of ultrasound. Uterine fibroids may in fact be an early warning sign of obstetrical complications with long-term impacts for maternal and child health.

## Data Availability

The datasets generated during and/or analyzed during the current study are not publicly
available due to Human Subject Protection requirements but are available from the
corresponding author on reasonable request and after IRB review and approval.

---

**Supplementary Table 1.** Clinical uterine fibroids diagnosis method.

| **Supplementary Table 1. Clinical uterine fibroids diagnosis method** |  |  |
| --- | --- | --- |
|  | **ICD 9** | **ICD 10** |
| Fibroids | 218, 218.0, 218.1, 218.2, 218.9 | D25, D25.0, D25.1, D 25.2, D25.9 |

**Supplementary Table 2.**
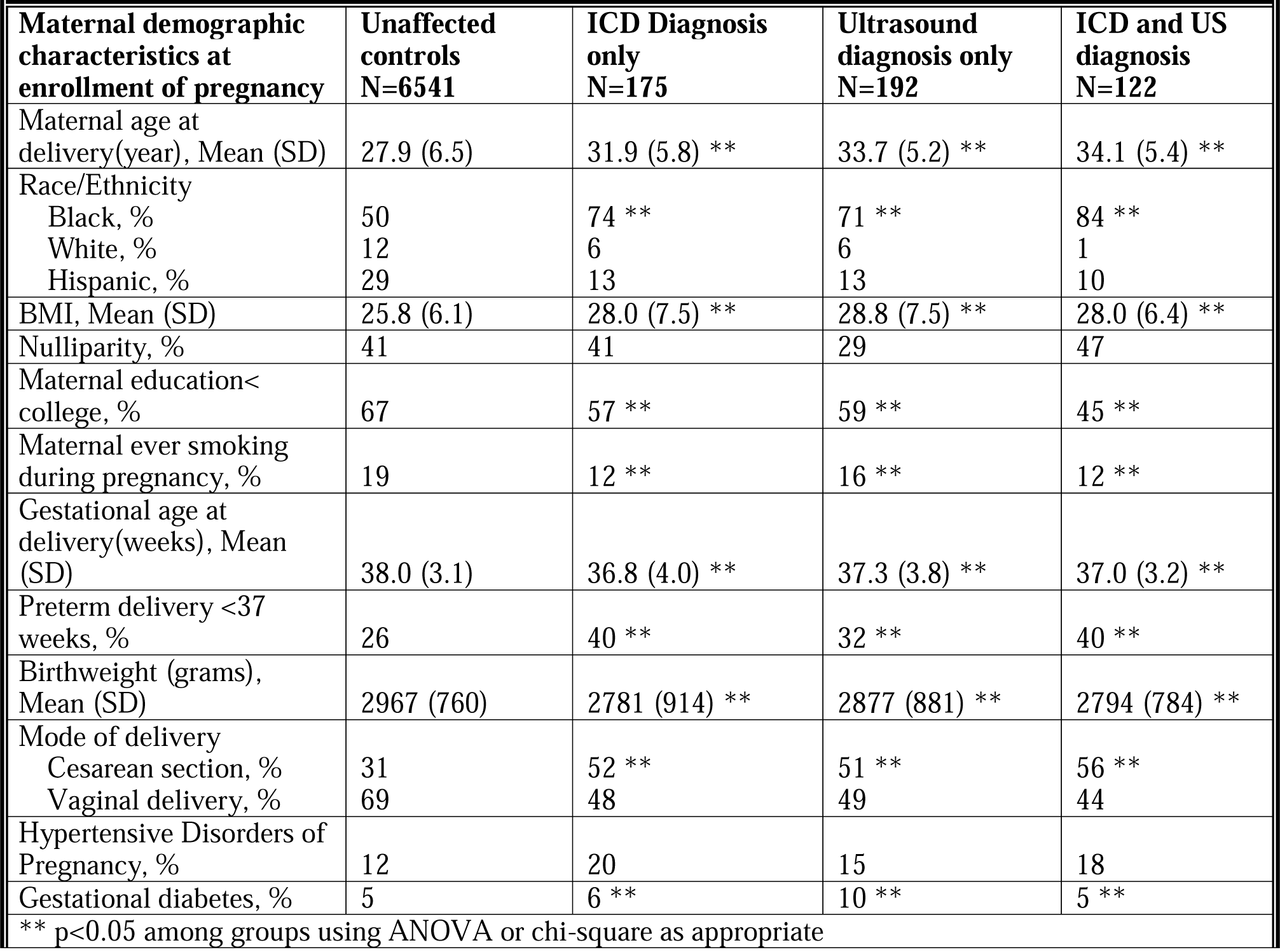
Demographic Characteristics of nested cohort mother-child dyads, BBC, based on fibroid diagnosis type.

## References

ACOG Practice Bulletin No. 202: Gestational Hypertension and Preeclampsia. Obstet Gynecol 2019;133:1.

Alsnes IV, Vatten LJ, Fraser A, Bjørngaard JH, Rich-Edwards J, Romundstad PR, Åsvold BO. Hypertension in Pregnancy and Offspring Cardiovascular Risk in Young Adulthood: Prospective and Sibling Studies in the HUNT Study (Nord-Trøndelag Health Study) in Norway. Hypertension 2017;69:591–598.

Biderman-Madar T, Sheiner E, Levy A, Potashnik G, Mazor M. Uterine leiomyoma among women who conceived following fertility treatment. Arch Gynecol Obstet 2005;272:218–222.

Boyd HA, Basit S, Behrens I, Leirgul E, Bundgaard H, Wohlfahrt J, Melbye M, Øyen N. Association Between Fetal Congenital Heart Defects and Maternal Risk of Hypertensive Disorders of Pregnancy in the Same Pregnancy and Across Pregnancies. Circulation 2017;136:39–48.

Boynton-Jarrett R, Rich-Edwards J, Malspeis S, Missmer SA, Wright R. A prospective study of hypertension and risk of uterine leiomyomata. Am J Epidemiol 2005;161:628–638.

Brewster LM, Haan Y, van Montfrans GA. Cardiometabolic Risk and Cardiovascular Disease in Young Women With Uterine Fibroids. Cureus 2022;14:e30740.

Chen Y, Lin M, Guo P, Xiao J, Huang X, Xu L, Xiong N, O’Gara MC, O’Meara M, Tan X. Uterine fibroids increase the risk of hypertensive disorders of pregnancy: a prospective cohort study. J Hypertens 2021;39:1002–1008.

Conti N, Tosti C, Pinzauti S, Tomaiuolo T, Cevenini G, Severi FM, Di Tommaso M, Petraglia F. Uterine fibroids affect pregnancy outcome in women over 30 years old: role of other risk factors. J Matern Fetal Neonatal Med 2013;26:584–587.

Coronado GD, Marshall LM, Schwartz SM. Complications in pregnancy, labor, and delivery with uterine leiomyomas: a population-based study. Obstet Gynecol 2000;95:764–769.

Farland LV, Prescott J, Sasamoto N, Tobias DK, Gaskins AJ, Stuart JJ, Carusi DA, Chavarro JE, Horne AW, Rich-Edwards J et al. Endometriosis and Risk of Adverse Pregnancy Outcomes. Obstet Gynecol 2019;134:527–536.

Girault A, Le Ray C, Chapron C, Goffinet F, Marcellin L. Leiomyomatous uterus and preterm birth: an exposed/unexposed monocentric cohort study. Am J Obstet Gynecol 2018;219:410.e1–410.e7.

Gong L, Liu M, Shi H, Huang Y. Uterine fibroids are associated with increased risk of pre-eclampsia: A case-control study. Front Cardiovasc Med 2022;9:1011311.

Haan YC, Diemer FS, Van Der Woude L, Van Montfrans GA, Oehlers GP, Brewster LM. The risk of hypertension and cardiovascular disease in women with uterine fibroids. J Clin Hypertens (Greenwich) 2018;20:718–726.

Harlev A, Wainstock T, Walfisch A, Landau D, Sheiner E. Perinatal outcome and long-term pediatric morbidity of pregnancies with a fibroid uterus. Early Hum Dev 2019;129:33–37.

Humphrey JD. Mechanisms of arterial remodeling in hypertension: coupled roles of wall shear and intramural stress. Hypertension 2008;52:195–200.

Kjerulff KH, Langenberg P, Seidman JD, Stolley PD, Guzinski GM. Uterine leiomyomas. Racial differences in severity, symptoms and age at diagnosis. J Reprod Med 1996;41:483–490.

Kuwahara F, Kai H, Tokuda K, Kai M, Takeshita A, Egashira K, Imaizumi T. Transforming growth factor-beta function blocking prevents myocardial fibrosis and diastolic dysfunction in pressure-overloaded rats. Circulation 2002;106:130–135.

Lee SJ, Ko HS, Na S, Bae JY, Seong WJ, Kim JW, Shin J, Cho HJ, Choi GY, Kim J et al. Nationwide population-based cohort study of adverse obstetric outcomes in pregnancies with myoma or following myomectomy: retrospective cohort study. BMC Pregnancy Childbirth 2020;20:716–9.

Michels KA, Velez Edwards DR, Baird DD, Savitz DA, Hartmann KE. Uterine leiomyomata and cesarean birth risk: a prospective cohort with standardized imaging. Ann Epidemiol 2014;24:122–126.

Miyashita-Ishiwata M, El Sabeh M, Reschke LD, Afrin S, Borahay MA. Differential response to hypoxia in leiomyoma and myometrial cells. Life Sci 2022;290:120238.

Nahum Sacks K, Friger M, Shoham-Vardi I, Spiegel E, Sergienko R, Landau D, Sheiner E. Prenatal exposure to preeclampsia as an independent risk factor for long-term cardiovascular morbidity of the offspring. Pregnancy Hypertens 2018;13:181–186.

Pan L, Fu Z, Yin P, Chen D. Pre-existing medical disorders as risk factors for preeclampsia: an exploratory case-control study. Hypertens Pregnancy 2019;38:245–251.

Pearson C, Bartell T, Wang G, Hong X, Rusk SA, Fu L, Cerda S, Bustamante-Helfrich B, Kuohung W, Yarrington C et al. Boston Birth Cohort Profile: Rationale and Study Design. Precis Nutr 2022;1:e00011.

Segars JH, Parrott EC, Nagel JD, Guo XC, Gao X, Birnbaum LS, Pinn VW, Dixon D. Proceedings from the Third National Institutes of Health International Congress on Advances in Uterine Leiomyoma Research: comprehensive review, conference summary and future recommendations. Hum Reprod Update 2014;20:309–333.

Stewart EA, Borah BJ. Uterine Fibroids and Hypertension: Steps Toward Understanding the Link. J Clin Endocrinol Metab 2021;106:e1039–e1041.

Stout MJ, Odibo AO, Graseck AS, Macones GA, Crane JP, Cahill AG. Leiomyomas at routine second-trimester ultrasound examination and adverse obstetric outcomes. Obstet Gynecol 2010;116:1056–1063.

Uimari O, Auvinen J, Jokelainen J, Puukka K, Ruokonen A, Järvelin M, Piltonen T, Keinänen-Kiukaanniemi S, Zondervan K, Järvelä I et al. Uterine fibroids and cardiovascular risk. Hum Reprod 2016;31:2689–2703.

Ying W, Catov JM, Ouyang P. Hypertensive Disorders of Pregnancy and Future Maternal Cardiovascular Risk. J Am Heart Assoc 2018;7:e009382.

Zhao SK, Wu P, Jones SH, Torstenson ES, Hartmann KE, Velez Edwards DR. Association of uterine fibroids with birthweight and gestational age. Ann Epidemiol 2020;50:35–40.e2.

